# Medical nurses may be effective in using silver diamine fluoride to prevent caries compared to dental hygienists in a school-based oral health program

**DOI:** 10.1101/2022.05.09.22274845

**Authors:** Ryan Richard Ruff, Tamarinda J. Barry Godín, Richard Niederman

## Abstract

**Background:** The sustainability of school-based oral health programs depends on the utilization of effective, efficient treatments and the availability of a trained clinical workforce. The objective of this study was to determine whether registered nurses are comparable to dental hygienists in the application and effectiveness of silver diamine fluoride (SDF) for the prevention of dental caries.

**Methods:** CariedAway was a school-based study of SDF versus dental sealants and atraumatic restorations. Within the SDF arm, subjects were treated by either a licensed dental hygienist or a registered nurse, both under the supervision of a pediatric dentist. Although initial treatment assignment in CariedAway was randomized, assignment to provider was not. The proportion of children who remained caries free after two years was assessed for non-inferiority using two-group proportion tests, adjusting for the clustering effect of schools.

**Results:** 417 children were analyzed including 298 treated by hygienists and 119 by nurses. The proportion of caries-free individuals was 0.812 and 0.798 for hygienists and nurses, respectively, for a difference of 0.014 (95% CI = -0.07, 0.098) and within the pre-determined non-inferiority margin.

**Conclusions:** Nurses may be effective in treating children with silver diamine fluoride in school-based oral health programs.

## Introduction

The World Health Organization estimates dental caries to be the world’s most common noncommunicable disease, disproportionately affecting low-income and minority populations and consuming between 5-10% of healthcare budgets of industrialized nations [1, 2]. Notably, those most at risk of dental caries typically lack access to traditional dental services, which results in substantial unmet need in vulnerable groups [3, 4, 5]. As a public health intervention to increase access to dental care, the Centers for Disease Control and Prevention recommends school-based sealant programs, which are effective [6, 7, 8], economical [9], and “allow the use of dental personnel to the top of their licensure” [10].

Silver diamine fluoride (SDF) is an efficient, economical treatment for dental caries [11] that is supported by the American Academy of Pediatric Dentists as part of a comprehensive caries management program [12]. Systematic reviews conclude that SDF is effective in the arrest of caries in primary teeth [13] and significantly reduces the development of new dentin caries after twenty-four months [14]. SDF is a colorless liquid similar in application to fluoride varnish [15], and the Association of State & Territorial Dental Directors (ASTDD) advocate for physicians, nurses, and assistants to provide SDF in addition to dentists and dental hygienists [16].

The CariedAway study was a longitudinal, school-based study of non-surgical interventions for dental caries [17]. Primary objectives of CariedAway were to compare the incidence and arrest of dental caries in children treated with either silver diamine fluoride or glass ionomer sealants and atraumatic restorations [18], and to assess oral health-related quality of life [19]. Within the SDF treatment arm, care was provided by either dental hygienists or registered medical nurses. Subjects were not randomized to different providers within this group. A secondary objective of CariedAway was to conduct a preliminary investigation of the variation in provider effectiveness in the use of SDF to prevent dental caries.

## Methods

CariedAway is a registered study at www.clinicaltrials.gov (NCT03442309) beginning 22 February 2018, and received ethical approval from the New York University School of Medicine IRB.

### Design and Participants

CariedAway was a longitudinal pragmatic non-inferiority trial conducted from 2018-2023 in New York City primary schools. The pragmatic design was chosen to test study hypotheses in real world settings that characterize school-based oral health programs. Schools that had a student population from at least 50% Hispanic/Latino or black ethnicities and at least 80% receiving free and reduced lunch (a proxy for low socio-economic status) were eligible for inclusion. Schools were further excluded if they already had a school-based oral health program that provided services. Within eligible and enrolled schools, all subjects were eligible for the study if they provided parental informed consent and child assent. Children were included in analysis if they were between the ages of 5 and 13 years.

### Randomization

Although subjects were randomized to receive silver diamine fluoride, they were not randomized to be treated by either a dental hygienist or registered medical nurse. No systematic efforts were made to assign subjects to different providers. By law, nurses could only treat patients if standing orders were created by a supervising dentist. Subjects who were enrolled prior to visiting the school were thus eligible to be seen by nurses or hygienists. These subjects were seen by whichever provider was available. Any subject enrolled during the actual visit to schools (e.g., day-of enrollment) were seen only by hygienists.

### Interventions and Data Collection

All subjects included in analysis received a single treatment consisting of 38% silver diamine fluoride (Elevate Oral Care Advantage Arrest 38%, 2.24 F-ion mg/dose) applied to all asymptomatic cavitated lesions and brushed on all pits and fissures of bicuspids and molars for 30 seconds. Fluoride varnish (5% NaF, Colgate PreviDent) was then applied to all teeth. Registered nurses and hygienists operated within a specific room in each school using a disposable mirror, disposable explorer, and head lamp with participants laying in a portable dental chair. Nurses and hygienists were under the supervision of a licensed pediatric dentist.

Study clinicians performed full-mouth visual-tactile oral examinations at each observation. Teeth were assessed as being present or missing intraorally. Individual tooth surfaces were assessed as being either intact/sound, sealed, restored, decayed, or arrested. All data were recorded on Apple iPads using an electronic dental health record designed for school-based programs (New England Survey Systems, Brookline, MA) and were securely uploaded each day to the Boston University School of Public Health Data Coordinating Center.

### Blinding

All clinicians in this analysis provided silver diamine fluoride and thus were not blinded to their treatment assignment. However, examiners at follow-up were unable to identify the type of clinician that provided initial treatment until the examination was completed.

### Clinician training

Registered nurses first completed modules on child oral health, caries risk assessment, fluoride varnish, and oral examinations from *Smiles for Life: A national oral health curriculum* [20], accredited by the American Nurses Credentialing Center’s Commission on Accreditation. Nurses and hygienists further received approximately 70 hours of didactic instruction and practical training during the annual CariedAway orientation, including dental screening and treatment protocols for silver diamine fluoride and fluoride varnish. Examiner standardization was conducted via case study, utilization of dental models, and through the recruitment of examinees attending pilot and training schools not included in the CariedAway study population. Examiners were standardized using identical diagnostic and treatment protocols. Caries diagnosis was performed according to guidelines of the International Caries Detection and Assessment System (ICDAS) adapted criteria in epidemiology and clinical research settings [21]. Full description of clinical protocols for diagnosis, outcomes, and other protocol considerations is previously published. Clinicians were standardized through agreement with the senior examiner, a licensed pediatric dentist experienced in applying the dental screening and treatment protocols. The senior examiner performed weekly chairside and data audits to ensure protocol compliance and continuous quality improvement.

### Impact of COVID-19

The original protocol for CariedAway stipulated that treatment and data collection were to be conducted biannually. However, the COVID-19 pandemic suspended medical interventions being conducted in New York City schools. As a result, the trial was conducted in two phases. In phase 1, we completed baseline observations and initial treatment from September 2019 to March 2020 with first follow-up observations completed from September 2021 to March 2022. In phase two, additional data collection was conducted in recurring six month intervals from March 2022 through June 2023.

### Outcomes

The primary outcome was the proportion of subjects with no observed incidence of decayed teeth from previously sound dentition (prevention). Nurses also provided treatment for existing caries at baseline, but these subjects were excluded from analysis as it was caries arrest.

### Covariates

Data for demographic variables were obtained from informed consent documents and/or school records, including race/ethnicity, age at observation, and sex. For socioeconomic status, all enrolled schools in CariedAway had at least 80% of the student population receiving free or reduced lunch (“Title 1” schools).

### Statistical Analysis

Analysis of provider variation used data from phase 1 of CariedAway.

Subjects were first ordered sequentially by visit and restricted to those assigned to the silver diamine fluoride arm and without dental caries at their baseline observation. Our primary independent variable, provider type, was dichotomized as either registered nurse or dental hygienist/pediatric dentist. Our primary dependent variable of any new evidence of dental decay was created as a dichotomous indicator reflecting whether each tooth presented at follow-up with either untreated caries or clinical evidence of having received an outside filling.

Non-inferiority of prevention by provider type was assessed using two-sample proportion tests with cluster adjustment for the school and the estimated intraclass correlation and by comparing the right-sided 95% confidence interval for the difference between providers to the pre-determined non-inferiority margin of 0.10. Power analysis for a two-group proportion test for non-inferiority with the predetermined threshold at 80% power was previously calculated to require a total sample size of 396. The degree of intraclass correlation was measured using intercept-only mixed effects multilevel regression modeling.

Results were subsequently compared to a confidence interval for the difference of hygienist minus nurse computed using bootstrap sampling with 10,000 replications. Logistic regression models were also conducted to explore the role of potential confounding variables on the provider/caries relationship.

Following per-protocol analysis, we conducted an intent to treat analysis using imputed data. Five datasets were generated for all subjects receiving silver diamine fluoride and not presenting with caries at baseline using multiple imputation and analyzed using logistic regression. Analysis was performed using Stata v16.0 and R v1.4. Confidence intervals were created at the 95% level and statistical significance was set at 0.05.

## Results

Of the 4718 subjects enrolled and randomized in Phase 1, 2998 were enrolled from viable grades. Those subjects enrolled in fourth and fifth grades were treated for ethical reasons but were not viable as they would age out of the study prior to follow-up. We completed follow-up data collection with 1398 subjects from September 2021 through 2 March 2022 (Figure 1). A total of 1070 enrolled subjects were treated with silver diamine fluoride and did not have caries at baseline, and follow-up data was collected in 418 subjects. One subject did not have provider recorded and thus was removed from analysis (N=417, Table 1). This analytic sample consisted of 298 subjects treated by dental hygienists and 119 treated by registered nurses. Approximately 10% of subjects presented at baseline with dental sealants. In keeping with the study inclusion criteria, 81% of subjects were of Hispanic/Latino or black ethnicity. The intraclass correlation coefficient was less than .00001. There were no adverse events reported in children treated by either nurses or hygienists.

**Table 1:** Demographics overall and by provider type for the baseline sample (N=1070) and follow-up sample (N=417)

|  | Baseline Sample |  |  |  |  |  | Follow-up Sample |  |  |  |  |  |
| --- | --- | --- | --- | --- | --- | --- | --- | --- | --- | --- | --- | --- |
|  | All |  | Hygienist/D<br>entist |  | Nurses |  | All |  | Hygienist/D<br>entist |  | Nurses |  |
|  | N/Me<br>an | % /<br>SD | N | % | N | % | N/Me<br>an | % /<br>SD | N | % | N | % |
| Subjects | 1070 | 100 | 755 | 100 | 315 | 100 | 417 | 100 | 298 | 71.46 | 119 | 28.54 |
| Female | 540 | 50.47 | 286 | 52.45 | 144 | 45.71 | 221 | 53 | 167 | 56.04 | 54 | 45.38 |
| Race |  |  |  |  |  |  |  |  |  |  |  |  |
| Hispanic | 481 | 44.95 | 372 | 49.27 | 109 | 34.6 | 240 | 57.55 | 176 | 59.06 | 64 | 53.78 |
| Black | 171 | 15.98 | 136 | 18.01 | 35 | 11.11 | 98 | 23.5 | 64 | 21.48 | 34 | 28.57 |
| White | 26 | 2.43 | 19 | 2.52 | 7 | 2.22 | 10 | 2.4 | 8 | 2.68 | 2 | 1.68 |
| Asian | 14 | 1.31 | 14 | 1.85 | 0 | 0 | 14 | 3.36 | 10 | 3.36 | 4 | 3.36 |
| Multiple | 21 | 1.96 | 15 | 1.99 | 6 | 1.9 | 5 | 1.2 | 4 | 1.34 | 1 | 0.84 |
| Other | 12 | 1.12 | 8 | 1.06 | 4 | 1.27 | 11 | 2.64 | 7 | 2.35 | 4 | 3.36 |
| DK/Missing | 345 | 32.24 | 191 | 25.29 | 154 | 48.89 | 39 | 9.35 | 29 | 9.73 | 10 | 8.4 |
| Untreated decay, follow-up | - | - | - | - | - | - | 61 | 14.43 | 46 | 15.44 | 15 | 12.61 |
| Sealants at baseline | 105 | 9.81 | 78 | 10.33 | 27 | 8.57 | 43 | 10.31 | 33 | 11.07 | 10 | 8.4 |

**Figure 1:**
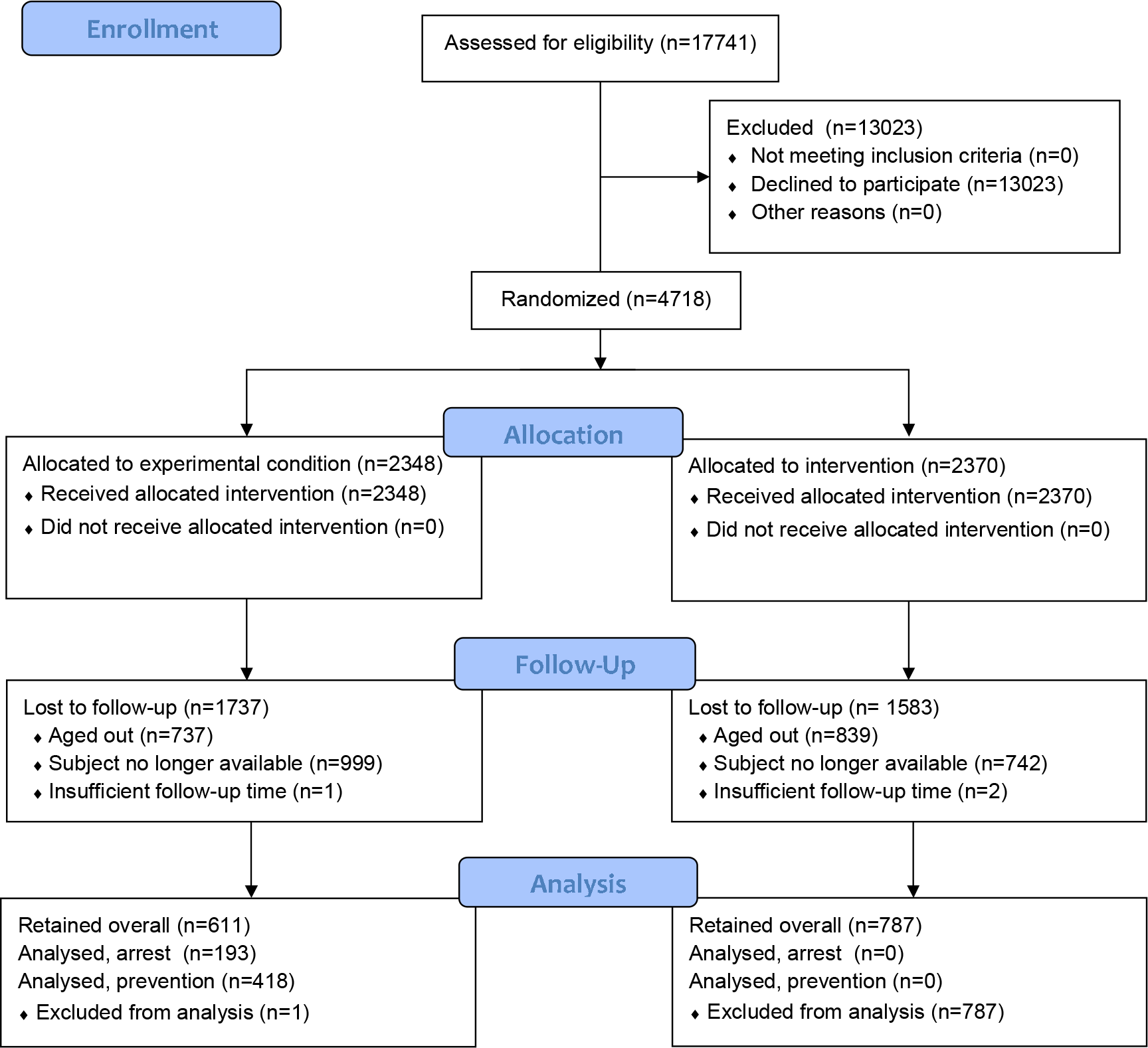
Study flow diagram for the CariedAway randomized controlled trial

The proportion of individuals who remained caries free after two years (prevention rate) was 0.812 and 0.798 for subjects treated by dental hygienists and nurses, respectively, for a difference of 0.014 (95% CI = -0.07, 0.098), just below the non-inferiority threshold (Table 2). Results were similar for bootstrapped confidence intervals (difference = 0.014, 95% CI = -0.09, 0.098). Results from regression models show no differences between provider after controlling for race, age at baseline, sex, and dental sealant prevalence (Table 3).

**Table 2:** Prevention rates after two years (N=417)

|  | Hygienists/Dentists |  |  | Nurses |  |  | Difference |  | 95% CI |  |
| --- | --- | --- | --- | --- | --- | --- | --- | --- | --- | --- |
|  | N | mean | SE | N | mean | SE | mean | SE | Lower | Upper |
| Prevention | 298 | 0.81 | 0.023 | 119 | 0.8 | 0.037 | 0.014 | 0.043 | -0.07 | 0.098 |

**Table 3:** Regression model results for provider type (N=417)

| Variable | OR | SE | 95% CIL | 95% CIU |
| --- | --- | --- | --- | --- |
| Provider | 0.9 | 0.25 | 0.52 | 1.55 |
| Race |  |  |  |  |
| Black | 0.78 | 0.23 | 0.43 | 1.41 |
| White | 0.52 | 0.37 | 0.13 | 2.1 |
| Asian | 1.35 | 1.06 | 0.29 | 6.29 |
| Other | 1.5 | 1.17 | 0.33 | 6.88 |
| Unreported | 0.63 | 0.26 | 0.28 | 1.4 |
| Age | 1.05 | 0.11 | 0.85 | 1.3 |
| Sex (male) | 1.19 | 0.3 | 0.72 | 1.96 |
| Sealants | 0.95 | 0.41 | 0.41 | 2.24 |

Imputed data for the baseline sample (N=1070) yielded 315 subjects with no baseline decay that were treated by nurses and 755 subjects by hygienists. The proportion of subjects who remained caries free at follow-up was 80.8% and 81% for nurses and hygienists, respectively. The odds ratio for prevention comparing nurses to hygienists was 1.01 (95% CI = 0.68, 1.50). Results with imputed data remained non-inferior.

## Discussion

Our results suggest that the effectiveness of silver diamine fluoride applied by registered nurses may be comparable in the two-year prevention of caries compared to dental hygienists in a school-based pragmatic setting. Specifically, the overall prevention rate in predominantly low-income, minority children was approximately 80%. If left untreated, dental caries can progress to severe infection [22] and negatively affects quality of life [23], academic performance [24], and school attendance [24]. Thus the effective prevention of dental caries in schools can not only improve health but overall child development.

The ubiquity and impact of school-based sealant programs depends in part on the availability of properly trained dental professionals. In a prior survey of registered dental hygienists, 54% of respondents were unfamiliar with SDF, 78% agreed that using SDF as a treatment for dental caries falls within their scope of practice, and 82% believed it to be an alternative to traditional restorative treatments [25]. Further, the use of traditional preventive services like glass ionomer sealants can be prohibitively expensive when treating large school populations, such as those found in New York City. In contrast, school nursing services prevent millions of dollars in medical care costs and lost productivity [26] and provide for safe and effective management of children with chronic health conditions, improving both health and academic outcomes [27]. State medical or dental practice acts can authorize nurses to treat children with silver diamine fluoride, thus their incorporation into school-based dental programs can have substantial impact on the efficiency and reach of care. For example, in New York, the state scope of practice implies that SDF is a topical fluoride and able to be applied by registered nurses under the supervision of a licensed dentist. This also aligns with professional practice, as the National Association of School Nurses advocates for the school nurse to promote child oral health through prevention, education, and coordination [28].

Our overall results are in line with the available evidence. Recent systematic reviews and meta-analyses are available for the effectiveness of silver diamine fluoride in the prevention of caries in primary dentition and permanent first molars [29, 14], caries control in exposed root surfaces [30], and in the arrest or reversal of both noncavitated and cavitated lesions in primary and permanent teeth [31]. These studies indicate preventive fractions for SDF versus placebo or active controls in the range of 70-90%, depending on the comparator used.

This study has multiple limitations. The provider assignment in the SDF arm of CariedAway was not a randomized one, and our results may be biased by unobserved differences across subjects. Children who enrolled prior to school visitation received standing orders from the supervising dentist and thus were able to be treated by either nurses or hygienists. These subjects were seen on a first come, first served basis and were not ordered by any factor such as disease severity, sex, or race. Those who enrolled during the study visit were only eligible to be treated by dental hygienist. If pre-visit enrollees were systematically different, this could introduce bias into our results. As a result, these findings should be interpreted with caution and considered as preliminary evidence. Further study with providers being a randomized factor is recommended.

The two year period between baseline and follow-up observations corresponded with the onset and duration of the COVID-19 pandemic, which resulted in the suspension of all in-school educational and clinical activities from March 2020 to August 2021. This had multiple potential implications. As our analytic sample considered only those with no untreated decay at baseline, exfoliation of decayed teeth was not a concern. Additionally, our analysis accounted for both any newly observed untreated decay and any evidence of having received a dental filling, which would suggest the incidence of new decay (and thus prevention failure) in the intervening years. As a result, our findings should not be confounded by receipt of traditional care in a dental office. Regardless, the low overall follow-up rate due to the effects of the COVID-19 pandemic remains a lingering concern, though our analysis with imputed subject data did not differ from primary results.

The CariedAway study utilized minimally-invasive treatments that were recently added to the World Health Organization’s list of essential medicines, which can be used to expand the scope and reach of school caries prevention. Our results support the incorporation of not only alternative non-restorative methods into traditional school-based prevention programs but the inclusion of under-utilized health professionals in the provision of care. As of 2018, an estimated 132,300 school nurses work in the United States [32], comprising approximately 75% of the entire hygienist workforce. This represents a substantial untapped resource to address oral health inequities.

## Data Availability

CariedAway is an ongoing clinical trial. Data are not publicly available. Data requests will be considered on a case by case basis.

## Author contributions

RRR and RN were the principal investigators and conceived of and designed the study. TBG served as the supervising dentist, directed clinical activities, and oversaw all data collection. RRR performed all statistical analyses and wrote the manuscript. All authors critically reviewed the manuscript and provided edits. All authors read and approved the final manuscript.

## Acknowledgements

We are grateful for the support of multiple people and organizations in their collaboration on this initiative, including Rachel Whittemore, Nydia Santiago-Galvin, and Denise Guerrero, without whom this work would not be possible. We are also thankful for our clinical team who provided care and our patient/stakeholder board who provided community perspectives on COVID-19 and school guidance. We also wish to thank John Roberge and Michael Stanley at New England Survey Systems (NESS), who worked with us to continuously improve our ability to capture clinical data, and Joe Palmissano at the Boston University School of Public Health. We thank Drs. Roger Platt, Ramneet Kalra, and David Tepel at the New York City Department of Health and Mental Hygiene, who facilitated access to New York City schools. Finally, we thank Drs. Dionne Richardson and Michele Griguts at the New York State Department of Health, who provided guidance during COVID-19.

---

**Table 4:** Comparison of nurses to hygienists in caries prevention, imputed data (N=1070)

| Variable | OR | SE | p-value | 95% CIL | 95% CIU |
| --- | --- | --- | --- | --- | --- |
| Provider | 1.01 | 0.2 | 0.944 | 0.68 | 1.5 |

